# Objective Olfactory Evaluation of Self-reported Loss of Smell in a Case Series of 86 COVID-19 Patients

**DOI:** 10.1101/2020.05.03.20088526

**Authors:** Jerome R. Lechien, Pierre Cabaraux, Carlos M. Chiesa-Estomba, Mohamad Khalife, Stéphane Hans, Christian Calvo-Henriquez, Delphine Martiny, Fabrice Journe, Leigh Sowerby, Sven Saussez

## Abstract

**Objective:** To investigate olfactory dysfunction (OD) in patients with mild COVID-19 through patient-reported outcome questionnaires and objective psychophysical testing.

**Methods:** COVID-19 patients with self-reported sudden-onset OD were recruited. Epidemiological and clinical data were collected. Nasal complaints were evaluated with the sino-nasal outcome-22 (SNOT-22). Subjective olfactory and gustatory status was evaluated with the National Health and Nutrition Examination Survey (NHNES). Objective OD was evaluated using psychophysical tests.

**Results:** Eighty-six patients completed the study. The most common symptoms were fatigue (72.9%), headache (60.0%), nasal obstruction (58.6%) and postnasal drip (48.6%). Total loss of smell was self-reported by 61.4% of patients. Objective olfactory testings identified 41 anosmic (47.7%), 12 hyposmic (14.0%), and 33 normosmic (38.3%) patients. There was no correlation between the objective test results and subjective reports of nasal obstruction or postnasal drip.

**Conclusion:** A significant proportion of COVID-19 patients reporting OD do not have OD on objective testing.

## Introduction

Since the onset of the coronavirus disease 2019 (COVID-19) pandemic in Europe, many otolaryngologists have reported patients with a sudden loss of smell.^1,2^ Olfactory dysfunction is rapidly becoming a key symptom of COVID-19, with more than 66% of patients in Europe and U.S reporting some degree of hyposmia.^1,3–6^ The loss of smell has been reported to occur before (11.8%), after (65.4%) or at the same time (22.8%) as the onset of other general or otolaryngological symptoms.^1^ Knowledge around the relationship between olfactory dysfunction and COVID-19 is rapidly evolving. Recently, Yan *et al*. shown that anosmia seems to be associated with a milder clinical course in patients with COVID-19.^6^ Moein *et al*. suggested that 98% of 60 Iranian COVID-19 patients exhibited some olfactory dysfunction on objective testing; only 35% of these patients were aware of hyposmia/anosmia before testing.^7^ The nuances around olfaction in COVID-19 appear to be associated with different clinical parameters than other symptoms, and, consequently, warrant further investigation.

The objective of this study was to investigate the olfactory dysfunction of COVID-19 patients with subjective validated patient-reported outcome questionnaires and objective psychophysical testing.

## Methods

The study protocol was approved by the ethics committee of the Jules Bordet Institute (Central Ethics Committee, IJB-0M011-3137). Patients were invited to participate and informed consent was obtained once inclusion criteria were met.

### Setting

Adult patients with confirmed COVID-19 and self-reported sudden-onset olfactory dysfunction were recruited through a public call from the Department of Anatomy of the University of Mons (Mons, Belgium). To be included, patients had to be not currently hospitalized (mild-to-moderate patients). The diagnosis of COVID-19 infection was based on the WHO interim guidance and symptoms of disease [8]. Individuals with self-reported sudden olfactory dysfunction and a clinical history suggestive of COVID-19 were invited to participate. A nasopharyngeal swab was performed to identify severe acute respiratory coronavirus-2 (SARS-CoV-2) via reverse transcription polymerase chain reaction (RT-PCR) for patient with symptom duration<14 days. In case of negative RT-PCR, serology for IgG and IgM to SARS-CoV-2 was realized. For patients with symptom duration ≥ 14 days, physicians performed serology (Figure 1). Only patients with a RT-PCR positive test or with positive IgG or IgM were included. Patients with a history of olfactory dysfunction before the pandemic, history of nasal surgery, chronic rhinosinusitis, head & neck trauma, or degenerative neurological disease were excluded from the study.

### COVID-19 diagnosis

The RT-PCR was performed by an experienced microbiologist (D.M.) in the LHUB-ULB laboratory of Brussels (Laboratoire Hospitalier Universitaire de Bruxelles, Belgium). Viral RNA extraction was performed by m2000 mSample Preparation SystemDNA Kit (Abbott) using 1000µl manually lysed sample (700µl sample + 800µl lysis buffer from kit) eluted in 90µl elution buffer. A qRT-PCR internal control was added at each extraction. qRT-PCR was performed using 10µl of extracted sample in the RealStar®SARS-CoV-2 RT-PCR Kit from Altona-diagnostics with a cut-off set at 40 cycle threshold (Ct).

Patients with a negative RT-PCR benefited from a serological test (Zentech, University of Liege Lab, Liege, Belgium) to determine whether or not they have been exposed to SARS-Cov-2.

### Epidemiological & Clinical outcomes

To minimize the risk of exposure for study personnel, the clinical and epidemiological characteristics of patients were electronically collected via an online questionnaire developed with Professional Survey Monkey® (San Mateo, California, USA). Demographic data including gender, age and ethnicity, as well as patient comorbidities and medications were collected.

### General and Otolaryngological Symptoms

The following general and ear, nose, and throat symptoms were collected and rated (from 0=no symptom to 4=severe symptom): cough, chest pain, dyspnea, headache, fever, fatigue, loss of appetite, myalgia, arthralgia, nausea, vomiting, diarrhea, excessive sticky sputum, skin manifestations (urticaria), conjunctivitis, nasal obstruction, postnasal drip, rhinorrhea, sore throat, facial pain, ear pain, dysphagia, dysphonia and dysgeusia. Dysgeusia was defined as the impairment of salty, sweet, bitter and sour.

### Patient-reported outcome questionnaires

The impact of COVID-19 on sinonasal symptoms was evaluated through the French version of the sino-nasal outcome test-22 (SNOT-22),^9^ a validated patient-reported outcome questionnaire from the original U.S. 20-item version.^10^

The impact of olfactory dysfunction on quality of life was assessed through the short version of the Questionnaire of Olfactory Disorders-Negative Statements (sQOD-NS).^11^ sQOD-NS is a 7-item patient-reported outcome questionnaire. Patients rated the item proposition from 0 (agree) to 3 (disagree) with total score ranging from 0 (significant impact of olfactory dysfunction on QoL) to 21 (no impact on QoL). Authors used sQOD-NS for its ease of completion.

The olfactory and gustatory questions were based on the smell and taste component of the National Health and Nutrition Examination Survey (NHNES).^12^ NHNES is a population survey that continuously monitors the health of adult citizens in the U.S. through a nationally representative sample of 5,000 persons on a yearly basis.^12^ The questions have been selected to characterize the variation, timing and associated-symptoms of both olfactory and gustatory dysfunction.

### Psychophysical Olfactory Evaluation

The psychophysical olfactory evaluations were performed using the identification sniffin’ sticks test (Medisense, Groningen, The Netherlands), which is a validated objective test of olfactory dysfunction.^13^ A total of 16 scents were presented via a pen device to patients for 3 seconds followed by a forced choice from 4 given options with a total possible score of 16 points. According to the results, patients were classified as normosmic (score between 12-16), hyposmic (score between 9-11) or anosmic (score 8 or below).

### Statistical Analyses

Statistical analyses were performed using the Statistical Package for the Social Sciences for Windows (SPSS version 22,0; IBM Corp, Armonk, NY, USA). The relationship between clinical and olfactory outcomes was analyzed through non-parametric test using Spearman correlation for scale data, Khi2 test for ordinal data and Mann-Whitney or Kruskal-Wallis test for categorized data. We investigated all potential associations between nasal complaints (nasal obstruction, rhinorrhea, postnasal drip) and the occurrence of olfactory disorder (sniffin’stick test). A level of significance of p<0.05 was used.

## Results

A total of 86 patients were eligible and completed the study (Figure 1). There were 56 females (65.1%) and 30 males (34.9%). The mean age was 42 ± 12 years. The majority of patients were Caucasian. Reflux, asthma and allergic rhinitis were the most common comorbidities (Table 1). Non-smokers accounted for 90% of the cohort.

**Table 1:** Epidemiological Characteristics of Patients.

footnotes: The mean PCR cycle number inversely reflects the viral load, According to the threshold of our Lab, 29 patients were positive for COVID-19 10 days (mean) after the initial diagnosis. Abbreviations: GERD=gastroesophageal reflux disease; SD=standard deviation.
| Characteristic | All patients<br>(N=86) |
| --- | --- |
| Age |  |
| Mean (SD) – yo | 41.7 ± 11.8 |
| Gender (N - %) |  |
| Female | 56 (65.1) |
| Male | 30 (34.9) |
| Ethnicity (N - %) |  |
| Caucasian | 84 (97.7) |
| North African | 2 (2.3) |
| Addictions (N - %) |  |
| Non-smoker | 77 (89.5) |
| Mild smoker (1-10 cigarettes daily) | 7 (8.1) |
| Moderate smoker (11-20 cigarettes daily) | 1 (1.2) |
| Heavy smoker (>20 cigarettes daily) | 1 (1.2) |
| Allergic patients | 16 (18.6) |
| Comorbidities |  |
| GERD | 9 (10.5) |
| Asthma | 5 (5.8) |
| Allergic rhinitis | 5 (5.8) |
| Hypertension | 4 (4.7) |
| Hypothyroidism | 3 (3.5) |
| Psoriasis | 2 (2.4) |
| Depression | 2 (2.3) |
| Sarcoidosis | 1 (1.2) |
| Hemochromatosis | 1 (1.2) |
| Obstructive apnea syndrome | 1 (1.2) |
| Autoimmune disease | 0 (0) |
| Diabetes | 0 (0) |
| Renal failure | 0 (0) |
| Hepatic insufficiency | 0 (0) |
| Respiratory insufficiency | 0 (0) |
| Heart problems | 0 (0) |
| Neurological diseases | 0 (0) |

### Clinical outcomes based on the general questionnaire

The most common general symptoms developed over the clinical course were fatigue (72.9%), headache (60.0%), cough (48.6%) and myalgia (42.9%) (Table 2). Fever, defined as a body temperature >38C°, was only reported by 8.6% of patients. Asthenia was the most commonly reported severe general symptom. The most common otolaryngological symptoms were nasal obstruction (58.6%), postnasal drip (48.6%), and dysgeusia (47.1%). Dysgeusia was considered the most severe otolaryngological symptom by half of the surveyed patients (Table 3).

**Table 2:** Severity of General Symptoms developed over the Clinical Course of the Disease (Percent of patients).

footnotes: The symptoms severity was assessed with a 4-point scale (from no problem (0) to Severe problem (4)).
| General Symptoms | 0= No<br>Problem | 1=Very Mild<br>Problem | 2=Mild or<br>Slight<br>Problem | 3=Moderate<br>Problem | 4=Severe<br>Problem |
| --- | --- | --- | --- | --- | --- |
| Fever | 64 (91.4) | 5 (7.1) | 1 (1.4) | 0 (0) | 0 (0) |
| Cough | 36 (51.4) | 18 (25.7) | 12 (17.1) | 4 (5.7) | 0 (0) |
| Chest pain | 57 (81.4) | 6 (8.6) | 5 (7.1) | 1 (1.4) | 1 (1.4) |
| Loss of appetite | 43 (61.4) | 10 (14.3) | 8 (11.4) | 5 (7.1) | 4 (5.7) |
| Sticky Sputum | 52 (73.4) | 11 (15.7) | 4 (5.7) | 1 (1.4) | 2 (2.9) |
| Arthralgia | 48 (68.6) | 8 (11.4) | 5 (7.1) | 7 (10.0) | 2 (2.9) |
| Myalgia | 40 (57.1) | 19 (27.1) | 3 (4.3) | 6 (8.6) | 2 (2.9) |
| Diarrhea | 48 (68.9) | 15 (21.4) | 4 (5.7) | 2 (2.9) | 1 (1.4) |
| Abdominal pain | 58 (82.9) | 8 (11.4) | 4 (5.7) | 0 (0) | 0 (0) |
| Nausea/vomitting | 61 (87.1) | 7 (10.0) | 2 (2.9) | 0 (0) | 0 (0) |
| Headache | 28 (40.0) | 17 (24.3) | 14 (20.0) | 10 (14.3) | 1 (1.4) |
| Asthenia | 19 (27.1) | 17 (24.3) | 14 (20.0) | 13 (18.6) | 7 (10.0) |
| Urticaria | 61 (87.1) | 3 (4.3) | 4 (5.7) | 1 (1.4) | 1 (1.4) |
| Conjunctivitis | 52 (74.3) | 12 (17.1) | 2 (2.9) | 3 (4.3) | 1 (1.4) |

**Table 3:** Severity of Ear, Nose, and Throat Symptoms developed over the Clinical Course of the Disease (Percent of patients).

footnotes: The symptoms severity was assessed with a 4-point scale (from no problem (0) to Severe problem (4)).
| **Ear, nose & throat** | 0= No | 1=Very Mild | 2=Mild or Slight | 3=Moderate | 4=Severe |
| --- | --- | --- | --- | --- | --- |
| **Symptoms** | Problem | Problem | Problem | Problem | Problem |
| Nasal obstruction | 29 (38.6) | 23 (32.9) | 12 (17.1) | 5 (7.1) | 1 (1.4) |
| Rhinorrhea | 37 (50.0) | 19 (27.1) | 12 (17.1) | 2 (2.9) | 0 (0) |
| Postnasal drip | 36 (48.6) | 17 (24.3) | 12 (17.1) | 5 (7.1) | 0 (0) |
| Throat pain | 52 (72.9) | 12 (17.1) | 3 (4.3) | 3 (4.3) | 0 (0) |
| Facial pain | 55 (77.1) | 7 (10.0) | 7 (10.0) | 1 (1.4) | 0 (0) |
| Ear pain | 47 (65.7) | 19 (27.1) | 2 (2.9) | 2 (2.9) | 0 (0) |
| Dysphagia | 63 (88.6) | 2 (2.9) | 4 (5.7) | 1 (1.4) | 0 (0) |
| Dyspnea | 52 (72.9) | 12 (17.1) | 3 (4.3) | 3 (4.3) | 0 (0) |
| Dysphonia | 53 (75.7) | 12 (17.1) | 2 (2.9) | 2 (2.9) | 1 (1.4) |
| Dysgeusia | 37 (52.9) | 4 (5.7) | 1 (1.4) | 7 (10.0) | 21 (30.0) |

### Patient Reported Outcome Questionnaire of Olfactory & Gustatory Function

According to the NHNES questions, 61.4% of patients described their olfactory disorder as total loss of smell at the onset of the disease, while the remainder reported partial loss. Cacosmia and phantosmia occurred in 34% and 20% patients, respectively. The mean scores of SNOT-22 and sQOD-NS are reported in Table 4.

**Table 4:**
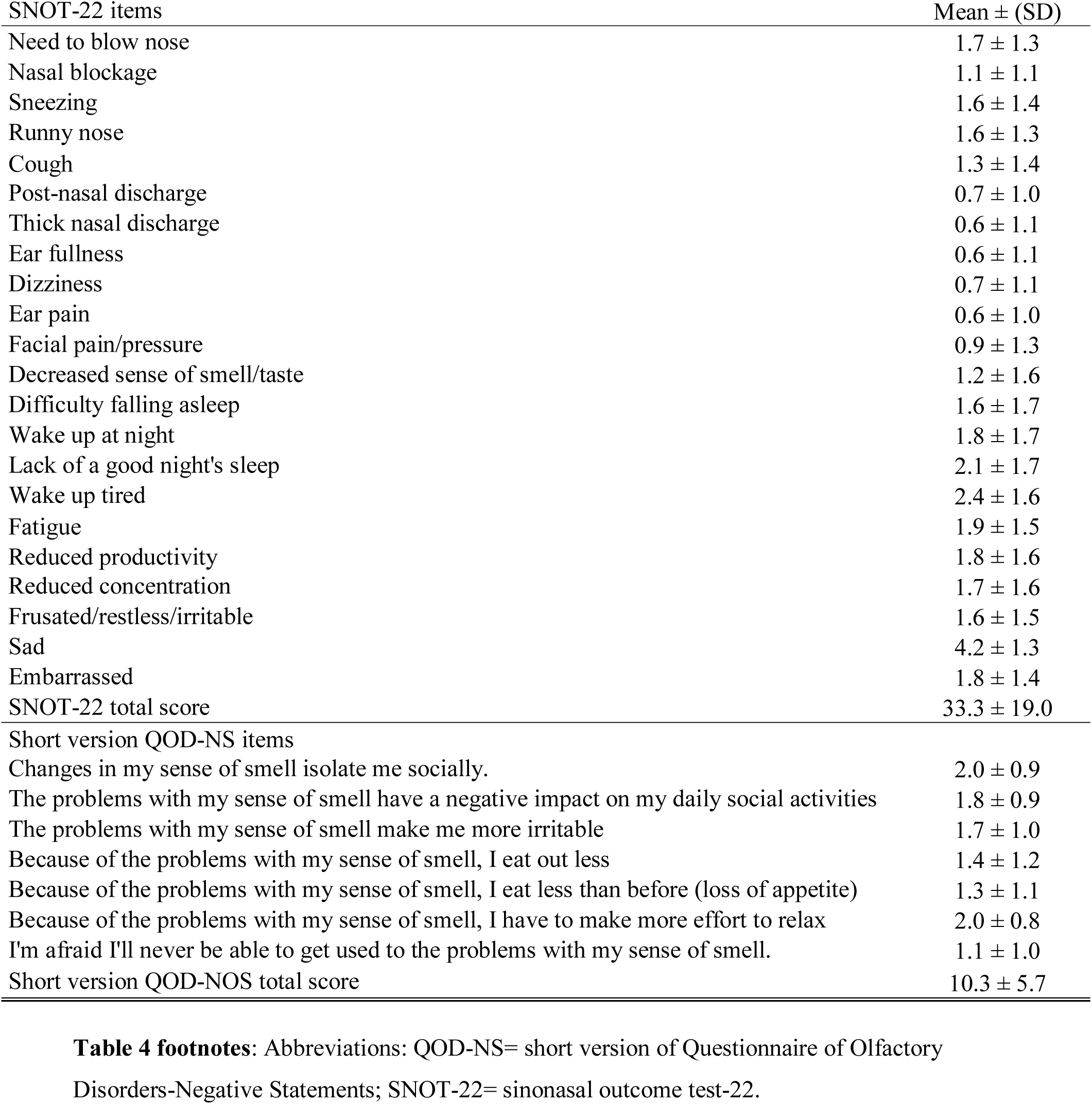
Sino-nasal Complaints of Patients with Olfactory Dysfunction.

Regarding gustatory dysfunction, 51% of patients reported taste disorders with abnormal sensations of salty, sweet, bitter and sour. The aroma perception was completely or partly lost in 42% and 32%, respectively, while 12% reported distortion of aroma.

### Psychophysical Olfactory Evaluations

The mean score of Sniffin’ Stick testing was 9 ± 4. Among the 86 patients, 41 (48%) and 12 (14%) patients were anosmic and hyposmic, respectively. A total of 33 (38%) patients who reported loss of smell were objectively normosmic. In the anosmic group, 26 (78.8%) patients reported total loss of smell. In the second group, 8 hyposmic individuals (88.9%) reported total loss of smell (Table 5).

**Table 5:** Characteristics of Anosmic, Normosmic and Hyposmic Patients.

footnotes: All 86 patients performed Sniffin'Stick tests.
|  | Anosmic (N=41) | Hyposmia (N=12) | Normosmia (N=33) | p-value | Test |
| --- | --- | --- | --- | --- | --- |
| Age – (Mean, SD) | 40 ± 12 | 39 ± 13 | 45 ± 11 | NS | KW |
| Sex (M/F) | 15/36 | 3/0 | 12/21 | NS | Khi2 |
| Tabacco (yes/no) | 5/36 | 2/10 | 2/31 | NS | Khi2 |
| Comorbidities (yes/no) |  |  |  |  |  |
| Hypertension | 0 | 2 | 2 | 0.027 | Khi2 |
| Rhinitis | 1 | 2 | 2 | NS | Khi2 |
| Reflux | 4 | 1 | 4 | NS | Khi2 |
| Asthma | 2 | 2 | 1 | NS | Khi2 |
| *SNOT-22 (Mean, SD)* | 33 ± 16 | 43 ± 20 | 34 ± 19 | NS | KW |
| Nasal obstruction (yes/no/NC) | 13/20/8 | 7/2/3 | 18/10/5 | NS | Khi2 |
| Self-reported Total loss of smell (N/%) | 26 (78.8) | 8 (88.9) | 18 (64.3) | NS |  |
| Duration of Anosmia (Mean, SD - days) | 17 ± 11 | 18 ± 11 | 17 ± 10 | NS | KW |

The mean durations of olfactory dysfunction at the time of the evaluations were 17 ± 11 days and 18 ± 11 days for anosmic and hyposmic patients, respectively. The mean duration of olfactory dysfunction of normosmic patients was 17 ± 11 days (Table 5).

Eleven patients realized sniffin’stick test twice (one week apart). Among these 11 patients, 9 were anosmic, 1 hyposmic and 1 normosmic at the first evaluation. From the first to the second visit (1 week later), the sniffin’stick test values improved in 5 patients (1 became hyposmic and 4 normosmic individuals) of the 9 anosmic patients of the first visit.

### Subgroup Analysis & Relationship between Outcomes

The nasal obstruction was not significantly associated with the development of olfactory dysfunction. Among the anosmic group, 60.1% of patients did not suffer from nasal obstruction (Table 5). There was no significant association between the results of the sniffin’ stick tests and the occurrence/severity of the following complaints: nasal obstruction and postnasal drip.

## Discussion

The involvement of COVID-19 in the development of olfactory and gustatory dysfunctions seems obvious. However, the characterization of the pathophysiological mechanisms underlying the olfactory dysfunction remains challenging regarding the risk of contamination. In this study, we have performed both subjective and objective olfactory evaluations in COVID-19 patients through online patient-reported outcome questionnaires and individual objective psychophysical testings. Interestingly, 38% of patients with self-reported olfactory dysfunction had normal olfactory testing at the sniffin’stick test.

The mismatch between the self-reported loss of smell and the anosmia regarding psychophysical testings has already been suggested in a recent Italian study where a few COVID-19 patients, who self-reported loss of smell, were objectively anosmic.^14^ Thus, the prevalence of olfactory dysfunction related to COVID-19 would be overestimated in the epidemiological studies where the loss of smell was based on subjective reports.

Another important finding of this study is the non-significant relationship between symptoms of nasal inflammation and objective olfactory dysfunction. In most cases of olfactory dysfunction occurring in viral infections, the olfactory disorder is related to the inflammatory reaction of the mucosa, leading to nasal obstruction, rhinorrhea and postnasal drip. In some cases, the olfactory dysfunction appeared to be related to other mechanisms, such as a neural spread of the virus into the neuroepithelium and the olfactory bulb. In 2007, Suzuki *et al*. demonstrated that coronavirus may be detected in the nasal discharge of patients with olfactory dysfunction.^15^ In this study, some patients had normal acoustic rhinometry, suggesting that nasal inflammation and related obstruction were not the only etiological factors underlying the olfactory dysfunction in viral infection. Netland *et al*. demonstrated on transgenic mice expressing the SARS-CoV receptor (human angiotensinconverting enzyme 2) that SARS-CoV may enter the brain through the olfactory bulb, leading to rapid transneuronal spread.^16^ The neurotropism of the COVID-19 is not new and would be associated with other symptoms and findings. For example, the virus spread into the central nervous system is currently suspected to play a key role in respiratory failure through an effect on the medullary cardiorespiratory center.^17^ Similarly, the existence of different patterns of gustatory and olfactory recoveries would be explained by selective neurological impairments.^1^ In other words, and suggested by the aroma and gustatory outcomes, the loss of taste would be not a retro-olfactory disorder in some patients. Future experimental and clinical studies are needed to better understand the pathophysiological mechanisms underlying the development of olfactory and gustatory dysfunctions. These studies would associate patient-reported outcome questionnaires, psychophysical olfactory evaluations, fiberoptic examinations, and imaging or neurophysiological assessments.

The main limitation of the present study is the heterogeneity between patients about the duration of the olfactory dysfunction. However, it is complicated to recruit patients at the first day of the olfactory disorder for many reasons. First, many patients have other troublesome symptoms (e.g. fatigue, myalgia, arthralgia), which may limit the realization of the tests. Second, the recruitment of patients at the first day of the olfactory dysfunction involved a continuous communication to recruit these patients. In practice, it is complicated to communicate with the general public every day for a scientific study. The lack of full objective methods to assess olfaction may be considered as another weakness. In this study, we decided to use the Identification sniffin’sticks test (16 items) for practical and ethical reasons. This test may be performed quickly, which is important to reduce the risk of potential contamination of caregivers.

## Conclusion

Only 62% of COVID-19 patients with self-reported olfactory dysfunction have anosmia or hyposmia on objective psychophysical olfactory evaluation. Interestingly, the majority of those with confirmed objective olfactory dysfunction did not have nasal inflammatory symptoms, supporting the need of future clinical and experimental studies to clarify the pathophysiological mechanisms underlying the development of anosmia in COVID-19.

## Data Availability

All data are available on demand.

---

**Figure 1**: Chart flow.

**Figure 1** footnote: Abbreviations: NHNES: National Health and Nutrition Examination Survey; SNOT-22: Sinonasal outcome tool-22; sQOD-NS: short version of Questionnaire of Olfactory Disorders-Negative Statements.

